# Observational Cross-Sectional Study to Estimate Population Norms in Eight Countries: the POPUP Study Protocol

**DOI:** 10.1101/2025.03.24.25324318

**Authors:** Sarah Dewilde, Nafthali Hananja Tollenaar, Glenn Phillips, Sandra Paci, Mathieu F. Janssen

## Abstract

This study protocol outlines the Population Norms Study (POPUP), a multinational digital survey aimed at establishing general population norms for medical resource use, comorbidities, sick leave, caregiver support, quality of life, and functioning across eight countries: United States (US), Canada, United Kingdom (UK), the Netherlands, Belgium, Spain, Italy, and Germany. Data will be collected through an online self-administered survey in two waves: the first in Q1 2021 and the second in Q1 2023, when first-wave responders will be recontacted. A total of 15,500 responses will be gathered across both waves: 9,000 in the first wave and 4,500 re-contacts in the second, with an additional 2,000 new contacts if needed. Representative panels will be recruited by a research company from each country based on age, gender, education, and region. Participants will complete the survey after providing informed consent. No personal identifiers will be recorded. The observational study involves no medical interventions, drugs, or devices, and will be submitted for Ethical Committee approval in each country. Descriptive statistics will be used for data analysis. The results are expected to provide a baseline for comparing health outcomes in specific patient populations and quantifying disease burden.

## 1. Background

Health-related quality of life (HRQoL), functioning, and the health and economic burden have become key outcomes in healthcare and public health interventions. Several instruments have been developed to assess these outcomes. Population norms for these measures serve as a baseline for comparing health outcomes in populations with specific health conditions, allowing for the quantification of disease burden. Methods used to obtain national or regional population norms include face-to-face interviews, computer-assisted personal interviews, postal surveys, computer-assisted telephone interviews, and, more recently, web surveys.^1^ Increasingly, patients who participate in studies fill out the quality of life instruments electronically.^2^ A review of papers studying the agreement between electronic and paper administration of patient-reported outcome measures in disease populations, found that the two modes of administration had a high level of equivalence.^3^ A Norwegian study compared a postal and a web survey to obtain general population norms for the EQ-5D-3L instrument. Results of this study supports the use of web surveys when collecting data for health-related quality of life population norms.^1^ National general population norms can be used as reference data to assess the burden of disease of patients with specific conditions.^4^

## 2. Objectives of the study

The objective of the Population Norms Study (POPUP) is to report general population norms for assessing medical resource use, comorbidities, sick leave, caregiver support, quality of life, and functioning across eight countries (US, Canada, UK, the Netherlands, Belgium, Spain, Italy, and Germany).

## 3. Study design

This observational multinational cross-sectional study that will collect data through an online self-administered survey in two waves. The first wave will be conducted in Q1 2021, and the second wave in Q1 2023, when first-wave responders were recontacted.

## 4. Study population

Representative panels of the general population from eight different countries (US, Canada, UK, the Netherlands, Belgium, Spain, Italy, and Germany) will be recruited. Participants will be recruited by a research company through online panels in each country. These panels are usually regularly updated, and low-quality respondents will be filtered out. Participants will be invited via email and incentivized with points redeemable for gifts.

## 5. Data collection

The following data will be collected from all panel participants. The first wave of data collection is expected to take one month. One year later, respondents will be contacted again for the second wave, which is also expected to take one month. To minimize missing data, the survey will be programmed to prevent skipped or unanswered questions. Licenses for the questionnaires will be purchased when applicable, and the surveys will be programmed using LimeSurvey. The time expected to complete the online questionnaire is 15 minutes.

### 5.1 Demographic data

Age, gender, level of education and current work situation.

### 5.2 Health and medical resource use related data

The presence of comorbidities and the number of days sick leave during the past month. The medical resources used during the past month:

- the need of a caregiver
- use of medical services (hospital, general practitioner, specialist)

### 5.3 Health-related quality of life and functioning

HRQoL and functioning will be measured using the following self-administered, validated instruments, each of which has been validated in the respective countries and languages:

➢ *Health Utilities Index 3* (HUI3) is a generic, preference-based, comprehensive system for measuring health status and health-related quality of life. The HUI3 classification system is comprised of eight attributes – Vision, Hearing, Speech, Ambulation, Dexterity, Emotion, Cognition and Pain – each with 5 or 6 levels of ability/disability. In this study the assessment period is current health and the assessment period the past 1-week.^5^
➢ *Myasthenia gravis activities of daily living* (MG-ADL) is an eight-question survey focussing on common symptoms of myasthenia gravis. Each item (respiratory function, the ability to brush teeth or comb hair, ability to rise from a chair, talking, chewing, vision and eyelid droop) is graded from 3 to 3.^6^
➢ *Myasthenia gravis quality of life measure, revised* (MG-QoLr) is a tool to assess aspects of life related to myasthenia gravis. The survey contains 15 items with 3 response options each.^7^
➢ *Hospital Anxiety and depression scale* (HADS) is a simple and reliable tool to assess anxiety and depression. Seven items in the questionnaire reflect depression and seven items anxiety. Each item has a 4-point response category.^8^
➢ *EQ-5D* is one of the most widely used instruments to describe and value health. The original questionnaire **(EQ-5D-3L)** contains five questions dealing with mobility, self-care, usual activities, pain/discomfort and anxiety/depression. All questions are answered on a scale ranging from 1 (best QoL) to 3 (worst QoL). The questionnaire contains a thermometer-like visual analogue scale (VAS) ranging from 0 to 100 (the EQ-VAS), where the endpoints are labelled ‘The best health you can imagine’ and ‘The worst health you can imagine’. The new **EQ-5D-5L** version expanded the response scale from three to five levels.^9^ **Bolt-ons** are dimension(s) that can be appended to another instrument. 6 bolt-ons (vision, breathing, sleep, tiredness, social relationships and self-confidence) in reference to EQ-5D-3L and EQ-5D-5L will be used in the survey.^10^

### 5.4 COVID-19 related questions

The impact of COVID-19 on the personal current situation will be evaluated through 7 short questions.

## 6. Sample size

The sample size for this observational study is not based on a formal calculation but is determined by the aim of ensuring representativeness of the general population and feasibility. However, a sample size of 1,000 provides a margin of error (ME) of 3.16% (ME% = 100 / √N), which is generally acceptable for surveys of the general population (**Table 1**).^11^ The total sample size consists of 15,500 responses across eight countries over two waves: 9,000 respondents in the first wave and 4,500 re-contacts in the second wave, supplemented with new contacts if the target sample size for the second wave is not reached. In all eight countries, samples will be drawn from representative panels based on age, gender, education, and region.

**Table 1.** POPUP Study Sample Size for Waves 1 and 2

| Country | Number of respondents in the first wave | Number of respondents in the second wave |
| --- | --- | --- |
| US | 2.000 | 1.000 |
| UK | 1.000 | 500 |
| Canada | 1.000 | 500 |
| Belgium | 1.000 | 500 |
| Spain | 1.000 | 500 |
| Italy | 1.000 | 500 |
| Germany | 1.000 | 500 |
| The Netherlands | 1.000 | 500 |
| **TOTAL SAMPLE SIZE** | **9.000** | **4500** |

## 7. Analyses

The study is purely descriptive. There is no formal hypothesis. Descriptive statistical methods will be used to analyse and describe the data. Pooled results for all the countries will be presented as well as results per country. The differences between countries will be explored. Results stratified by demographic characteristics such as age and gender will also be reported. Population norms for each of the research areas and instruments will be estimated for the different countries. It is planned to use the statistical analysis software SAS and R to analyse the collected data.

## 8. Procedures

### 8.1 Recruitment

Potential participants will be invited to participate through an e-mail invitation, with a link to the questionnaire. The sample of potential participants is aimed to be representative for the general population in each country. The sample will be drawn from a representative panel of survey participants willing to participate.

### 8.2 Data Quality Monitoring

Data in the e-platform will be checked for validity and internal consistency, to ensure high data quality and completeness.

### 8.3 Data protection

Names or other personal identifiers will not be recorded and therefore not be included in any database.

### 8.4 Funding

The research is sponsored by the pharmaceutical company Argenx BVBA.

### 8.5 Economic conditions

Individuals participating in the study will receive a small compensation for participating; a voucher or points for a lottery depending on the country.

### 8.6 Publication and data sharing policy

The results of this study will be published as individual papers. The publications will be aimed towards high impact peer-reviewed journals.

### 8.7 Timetable and end of the study

It is planned to start data collection in September 2020. The period needed to collect data from 4.500 individuals is estimated to be one month and will end when all data have been collected. It is aimed to obtain all data by December 2020.

### 8.8 Ethical considerations

Participants can only complete the online survey after agreeing to the informed consent form. No personal identifiers, including names, addresses, email addresses, phone numbers, postal codes, dates of birth, or national register numbers, will be recorded or included in any database. The participants are members of the general public, not individuals with a specific disease. This study is purely observational, with no medical interventions, drugs, or medical devices involved. The study will be submitted for Ethical Committee approval in each country.

## 9. Study organisation

### 9.1 Co-ordinating office

SHE BV, Boulevard Lambermont 418, 1030 Brussels, Belgium Mobile telephone: 0032 484 064 210.

## Data Availability

Anonymized, aggregated study data is available upon reasonable request through the corresponding author.

